# Functional networks of reward and punishment processing and their molecular profiles predicting the severity of young adult drinking

**DOI:** 10.1101/2024.02.06.24302417

**Authors:** Yashuang Li, Lin Yang, Dongmei Hao, Yu Chen, Bao Li, Youjun Liu, Yiyao Ye-Lin, Chiang-Shan R. Li, Guangfei Li

**Affiliations:** Department of Biomedical Engineering, College of Chemistry and Life Sciences, Beijing University of Technology, Beijing, China; Beijing International Science and Technology Cooperation Base for Intelligent Physiological Measurement and Clinical Transformation, Beijing, China; Department of Psychiatry, Yale University School of Medicine, New Haven, CT, USA; Centro de Investigación e Innovación en Bioingeniería, Universitat Politècnica de València, 46022 Valencia, Spain; Department of Neuroscience, Yale University School of Medicine, New Haven, CT, USA; Interdepartmental Neuroscience Program, Yale University School of Medicine, New Haven, CT, USA; Wu Tsai Institute, Yale University, New Haven, CT, USA (Running title: alcohol misuse and CPM)

**Keywords:** alcohol use disorder, alcohol misuse, fMRI, connectome, neurotransmitter, receptor

## Abstract

**Background:** Alcohol misuse is associated with altered punishment and reward processing. Here, we investigated neural network responses to reward and punishment and the molecular profiles of the connectivity features predicting alcohol use severity in young adults.

**Methods:** We curated the Human Connectome Project data and employed connectome-based predictive modeling (CPM) to examine how functional connectivity (FC) features during wins and losses associated with alcohol use severity in 981 young adults. Alcohol use severity was quantified by the first principal component of principal component analysis of all drinking measures of the Semi-Structured Assessment for the Genetics of Alcoholism. We combined the CPM findings and JuSpace toolbox to characterize the molecular profiles of the network connectivity features of alcohol use severity.

**Results:** The connectomics predicting alcohol use severity appeared specific, comprising less than 0.12% of all connectivity features. These connectivities featured the medial frontal, motor/sensory, and cerebellum/brainstem networks during punishment processing and medial frontal, fronto-parietal, and motor/sensory networks during reward processing. Spatial correlation analyses showed that these networks were associated predominantly with serotonergic and GABAa signaling.

**Conclusions:** A distinct pattern of network connectivity predicted alcohol use severity in young adult drinkers. These network features were associated with the serotonergic and GABAa signaling. These “neural fingerprints” help in elucidating the impact of alcohol misuse on the brain and providing evidence of new targets for future intervention.

## 1. Introduction

### 1.1 Reward and punishment processing in alcohol misuse

People may engage in drinking because of the positive (e.g., elevated sociability and physical relaxation) and/or the negative reinforcing (e.g., amelioration of negative emotions) effects of alcohol (Koob and Volkow, 2016). Distinguishing the mechanisms of positive and negative reinforcement helps in understanding of pathophysiology of craving (Yu Chen and Li, 2023) and how reward and punishment processing may interact with self-control in the etiological processes of alcohol misuse (Kahn et al., 2018).

Brain imaging provides a venue to elucidating the neural bases of altered reward and punishment processing in alcohol misuse. For instance, higher levels of reward sensitivity may contribute to alcohol misuse, as did lower ventral striatal activity during reward anticipation (Moreno Padilla et al., 2017). A study using both the Human Connectome Project (HCP) and Genetic Neuroimaging (IMAGEN) datasets reported higher functional connectivity of the medial orbitofrontal cortex, a reward area, and impulsivity in heavy drinkers (Cheng et al., 2019). In contrast, in our recent study, we demonstrated that loss rather than win reactivity along with fronto-striatal responses during a gambling task captured individual variation in alcohol use severity in a neurotypical sample (Li et al., 2022). In a reward go/no-go task where participants needed to initiate action or to inhibit an action to win money and/or avoid monetary loss, heightened punishment sensitivity enhanced the neural activities of avoidance and, in turn, contributed to alcohol misuse (Le et al., 2019). This literature together highlights the relevance of psychological and neural processes of reward and punishment processing to the pathophysiology of alcohol misuse.

### 1.2 Functional connectomics of individual traits and neuropsychiatric diagnoses

Connectome-based predictive modeling (CPM) is a data-driven approach for developing predictive models of brain behavior relationships and individual variability (Finn et al., 2015; Shen et al., 2017). The model also allows “computational lesion” to reveal features that are important in prediction (Feng et al., 2018). With CPM investigators extracted and summarized the most relevant connectivity features that were cross-validated in test data, and provided an estimate of the accuracy at which these features predict individual traits, including fluid intelligence (Finn et al., 2015), sustained attention (Rosenberg et al., 2016), mean sleep duration (Mummaneni et al., 2023), creative ability (Beaty et al., 2018), and drug craving (Antons et al., 2023), or clinical conditions, including akinetic rigidity of Parkinson’s disease (Wu et al., 2023) and binge drinking (Tong et al., 2021). For instance, with longitudinal multisite functional magnetic resonance imaging (fMRI) data collected at ages 14 and 19 to assess whole-brain patterns of functional organization that predict alcohol use, a recent study identified networks associated with vulnerability for future and current problem drinking (Antons et al., 2023). These studies of CPM characterized the systems-level neural markers that predict individual variation in health and illness.

### 1.3 Molecular profiles of functional brain networks

A critical approach to complementing systems-level findings is to investigate the molecular profiles of the neural networks identified of MRI. A tool for spatial correlation analyses of MRI data with nuclear imaging derived neurotransmitter maps, JuSpace provides a biologically meaningful framework to this end (Dukart et al., 2021) and offers novel insight into disease mechanisms and associated clinical features (Premi et al., 2023). For instance, in a study of frontotemporal dementia, investigators associated the altered patterns of grey matter volume (GMV) with the distribution of dopamine and acetylcholine pathways in mutation carriers and showed more widespread involvement of dopamine, serotonin, glutamate and acetylcholine pathways in symptomatic individuals (Pengo et al., 2023). Another work focused on a population of unmedicated first-episode schizophrenia and reported GMV alterations in association with the expression of serotonin, dopamine, and gamma amino butyric acid (GABA) receptors and/or transporters (Jingli Chen et al., 2023). Other studies combined MRI findings and JuSpace mapping to investigate the patterns of volumetric atrophy in link with changes in neurotransmitter pathways in Parkinson’s disease (Ren et al., 2023), multiple sclerosis (Fiore et al., 2023), and primary progressive aphasia (Premi et al., 2023).

Investigators have also combined multimodal brain imaging with JuSpace to link systems and molecular findings. Using functional connectivity density mapping of resting-state fMRI data along with a Go/No-Go task (outside scanner), Cui and colleagues reported spatial correlation with the ability of behavioral inhibition in the patterns of expression of gene categories involving cellular and synaptic elements of the cerebral cortex and ion channel activity as well as the serotonergic system (Cui et al., 2023). An earlier study employed both resting-state fMRI and structural MRI and associated changes in GMV and intrinsic connectivities with serotonergic, dopaminergic and μ-opioid receptor systems in heavy cannabis users (Hirjak et al., 2022). Tang and colleagues investigated volumetric atrophy, glucose hypometabolism, and neurotransmitter distribution utilizing both MRI and positron emission tomography data in Rasmussen’s encephalitis (Tang et al., 2022). Another study conducted meta-analytic co-activation analyses on lesion masks of individuals who acquired antisocial behaviors following their brain lesions and implicated multiple cortical and subcortical areas as well as the serotoninergic system (Dugre and Potvin, 2022).

Together, this literature supports the combination of MRI and JuSpace tools to associate systems and molecular profiles of cerebral dysfunction to better understand the pathophysiology of neurological and psychiatric conditions, including alcohol misuse.

### 1.4 The present study

Here, we applied CPM to fMRI data of a gambling task obtained from the Human Connectome Project (HCP). We characterized the connectivity features that predicted drinking severity. The gambling task involved win (reward) and loss (punishment) processing, and our previous study highlighted loss and fronto-striatal reactivities more than win reactivities in distinguishing individual severity of alcohol use (Li et al., 2022). In our first aim, we tested the hypotheses that whole-brain connectivity features during loss as compared to win processing would likewise better characterize alcohol use severity in this HCP sample of young adults. In our second aim, we combined the CPM findings and the JuSpace toolbox to investigate how the maps of connectivity features of alcohol use severity related to the molecular profiles and to better understand the molecular and cellular mechanisms underlying alcohol misuse.

## 2 Materials and Methods

### 2.1 Dataset and demographics

With permission from the HCP (Van Essen et al., 2012) and as in our previous work (Ide et al., 2020; Li et al., 2020; Peng et al., 2020; Li et al., 2021a; Li et al., 2021b; Li et al., 2022), we employed the 1200 Subjects Release (S1200) data set, which includes 3T MR imaging and behavioral data collected of the gambling task from 1080 subjects. A total of 981 subjects (473 men; mean ± SD = 27.9 ± 3.6 years; 508 women, 29.6 ± 3.6 years) were included in this study, after exclusion of 99 with head movements > 2 mm in translation or 2 degrees in rotation or for whom the images failed in registration to the template. All subjects were physically healthy with no severe neurodevelopmental, neuropsychiatric or neurological disorders. Because men and women differed significantly in age, age and sex were included as covariates in all analyses. The study was carried out in accordance with the latest version of the Declaration of Helsinki. HCP was approved by the Washington University Institutional Review Board (IRB #201204036).

### 2.2 Clinical measures

We assessed drinking severity based on 15 intrinsically associated measures of alcohol consumption in the past year, as evaluated by the Semi-Structured Assessment for the Genetics of Alcoholism. With principal component analysis (PCA) to reduce the dimensionality of the 15 parameters, we used the first principal component or PC1 as a quantitative index of alcohol use severity. It should be noted that some of the 15 measures needed to be flipped in sign to reflect severity of drinking. PC1 but no other PC’s had an eigenvalue > 1 and accounted for 60.73% of the variance of the data.

### 2.3 Neuroimaging data acquisition

Participants completed two runs of a gambling task each with 4 blocks (∼3 m and 12 s each run) – 2 each of reward and punishment, each with more win than loss trials and more loss than win trials – and a fixation period (baseline, 15 s) between blocks (Barch et al., 2013). Preprocessing and plotting was conducted using SPM8 and the BioImage Suite, as described in the **Supplement**.

### 2.4 Functional connectivity and connectome-based predictive modeling (CPM)

Whole-brain functional connectivity analyses were conducted using the BioImage suite. Network nodes were defined using the Shen 268-node brain atlas, which includes the cortex, subcortex and cerebellum. Task connectivity was calculated based on the ‘raw’ task time courses (punishment block only or reward block only). This involved computation of mean time courses for each of the 268 nodes (i.e., average time course of voxels within the node) for use in node-by-node pairwise Pearson’s correlations. R values of the 268×268 connectivity matrices represented the strength of connection between two individual nodes.

CPM was conducted using validated custom MATLAB scripts (Shen et al., 2017). CPM took group connectivity matrices and behavioral data (in this case PC1) as inputs to generate a predictive model of PC1 from connectivity matrices. Edges and PC1 from the training dataset were correlated using regression analyses to identify positive and negative predictive networks. Positive and negative networks were networks for which higher and lower edge weights (connectivity), respectively, were associated with PC1. While both networks were used for predicting PC1, they were by definition independent as a single edge could not be both a positive and negative predictor. Single subject summary statistics were then created as the sum of the significant edge weights in each network and entered into predictive models assuming linear relationships with PC1. The resultant polynomial coefficients (linear equation including slope and intercept) were then applied to the test dataset to predict PC1. We employed leave-one-out cross-validation, where a single “left-out” participant’s predicted value was generated by taking the data from all other participants as the training dataset in an iterative manner until all participants had a predicted value. Model performance (i.e., correspondence between predicted and actual values) was assessed using Spearman’s rho correlations. In leave-one-out cross-validation, analyses in the leave-one-out folds were not truly independent and the number of degrees of freedom was thus overestimated for parametric p-values of correlation. Instead of parametric testing, we therefore performed permutation testing. To generate the null distributions for significance testing, we randomly shuffled the correspondence between PC1’s and connectivity matrices 1,000 times and re-ran the CPM analysis with the shuffled data. Based on the null distributions, the p-values for leave-one-out predictions were calculated.

### 2.5 Correlation with neurotransmitters

JuSpace (https://github.com/juryxy/JuSpace) allows for spatial correlation analyses between cross-modal neuroimaging data (Dukart et al., 2021). JuSpace consists of a group of Matlab functions together with PET imaging maps of various receptor and transporter systems, each with a distinct atlas. All receptor and transporter maps were derived of an average of 6 to 174 healthy volunteers and linearly rescaled to a minimum of 0 and a maximum of 100. To determine the neurochemical basis underlying the neural networks of drinking severity, we computed the spatial correlations of the SPM T maps derived from whole brain regression each of “punishment-baseline” and “reward-baseline” against drinking PC1 and JuSpace maps of serotonin receptor (including 5-HT1a_1, 5-HT1a_2, 5-HT1b_1, 5-HT1b_2, 5-HT2a_1, 5-HT2a_2, 5-HT4); cannabinoid type I receptor (CB1); dopamine receptor (including D1,D2_1,D2_2); dopamine synthesis capacity receptor (FDOPA); gamma-aminobutric acid receptor (including GABAa_1, GABAa_2); mu opioid receptor (including MOR_1,MOR_2); metabotropic glutamate receptor (including mGluR5_1, mGluR5_2, mGluR5_3); dopamine transporter (DAT); noradrenaline transporter (NAT); serotonin transporter (including SERT_1, SERT_2, SERT_3); vesicular acetylcholine transporter (including VAChT_1, VAChT_2, VAChT_3). Pearson correlation coefficients between the T map and these neurotransmitter maps were calculated across the 119 brain regions of the neuromorphometrics atlas in JuSpace (Dukart et al., 2021) excluding all white matter and cerebrospinal fluid regions. Pearson’s correlation with p < 0.05 were considered significant.

## 3. Results

### 3.1 Predicting drinking severity PC1: loss processing

#### 3.1.1 CPM of loss processing

In CPM of loss-related processing in the gambling task (**Figure 1A**), the connectomics (positive and negative networks combined) successfully predicted drinking severity PC1 (r = 0.25, *p* = 0.001), as did connectivity within the positive (r = 0.24, *p* = 0.001) and negative (r = 0.25, *p* = 0.001) networks separately (**Figure 1B**).

**Figure 1.**
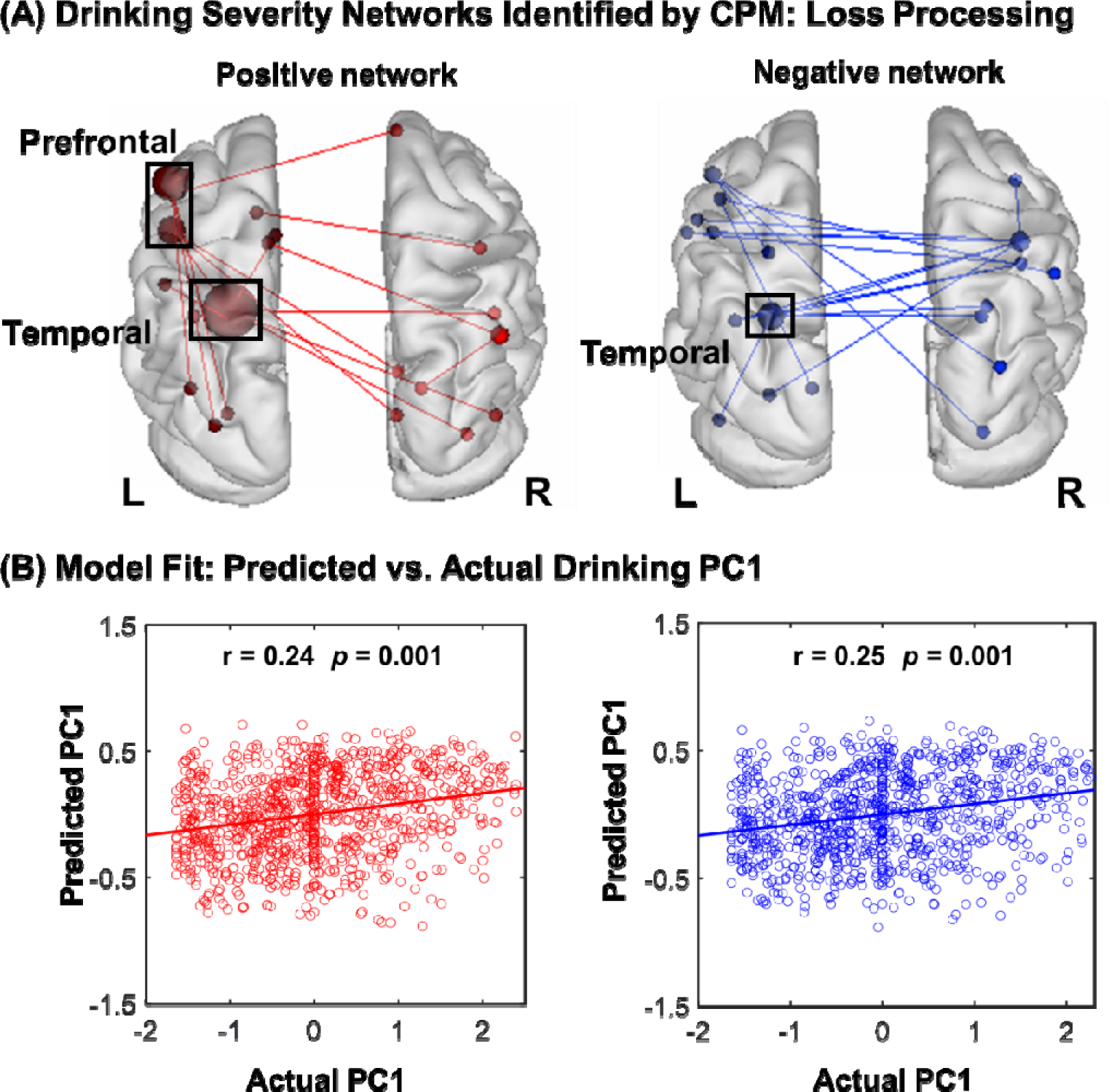
Macroscale network connectivities of loss processing in predicting drinking PC1. **(A)** shows positive (red) and negative (blue) networks in correlation with PC1. Larger spheres indicate nodes with more edges, and smaller spheres indicate fewer edges. For the positive network, higher edge weights (i.e., connectivity) predicted more severe drinking. For the negative network, lower edge weights predicted more severe drinking. **(B)** illustrates the correlation between actual (x-axis) and predicted (y-axis) drinking PC1 values generated using CPM.

#### 3.1.2 Network anatomy of loss processing

We summarized positive and negative networks based on the connectivities between macroscale brain regions (**Figure 2A**). Note that brain regions are presented in approximate anatomical order, such that longer-range connections are represented by longer lines. The network anatomies were complex and included connections between frontal, temporal, parietal lobes, cerebellum, and brainstem. Despite this complexity, these networks appeared to be quite specific, with positive and negative networks together including only 39 edges (17 positive, 22 negative), or less than 0.11% of all possible connections correlated with drinking severity PC1. Highest-degree nodes (i.e., nodes with the most connections) for the positive network included a temporal node with connections to prefrontal, parietal, limbic, cerebellar and other temporal nodes and prefrontal nodes with connections to occipital and temporal cortex. Highest-degree nodes for the negative network also included a temporal node with connections to the insula, limbic nodes as well as with connections to cerebellar and other temporal nodes. Both networks included short- and long-range connections. All edges showing significant correlation with PC1 are shown in **Supplementary Table S1**.

**Figure 2.**
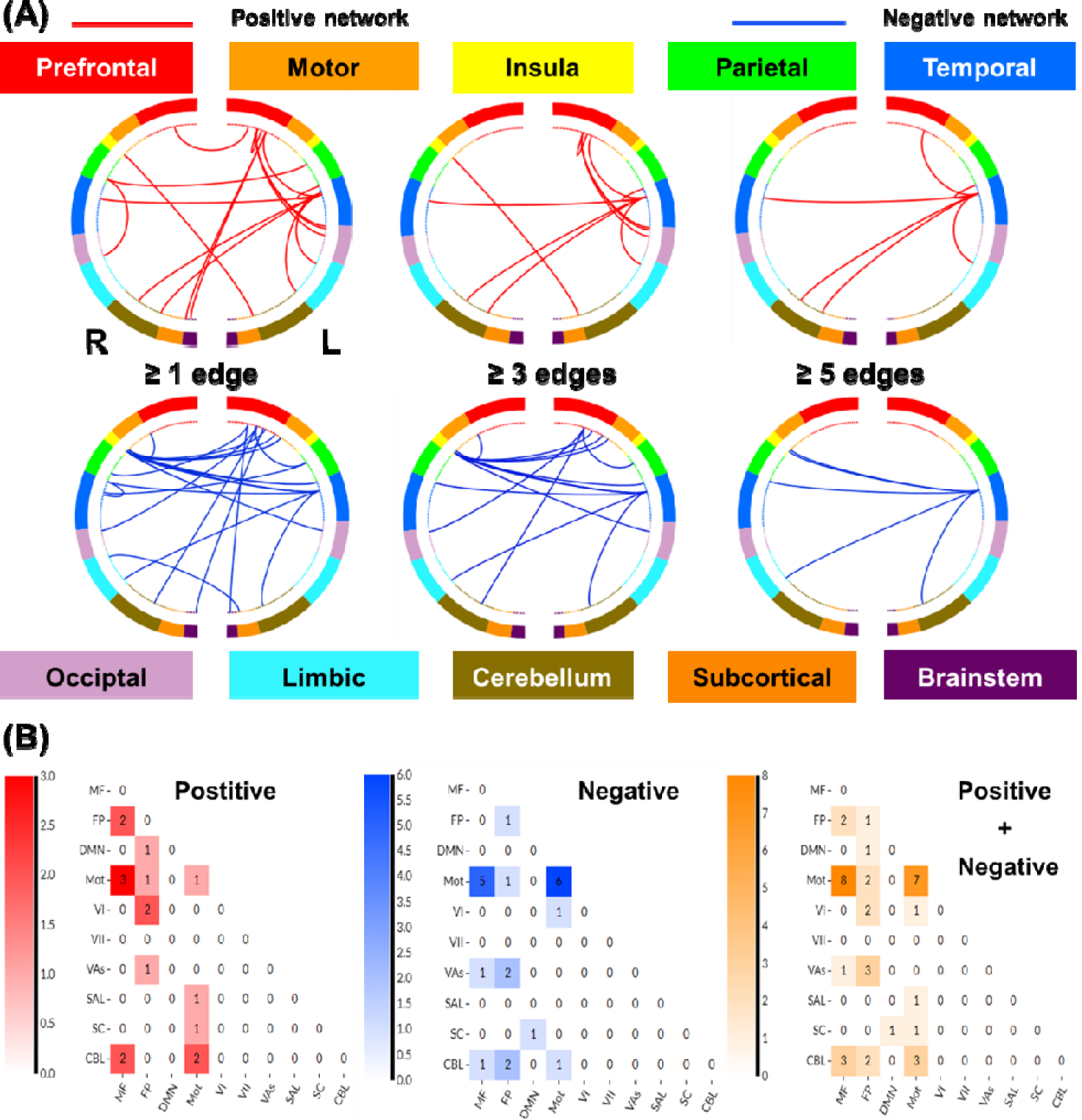
Macroscale network connectivities of loss processing in predicting drinking PC1. **(A)** Positive and negative networks summarized by connectivity between macroscale brain regions during punishment blocks. From the top, brain regions are presented in approximate anatomical order, such that longer-range connections are represented by longer lines. **(B)** Positive and negative and the sum of positive and negative networks. Cells represent the total number of edges connecting nodes within and between each network, with a higher number indicating a greater number of edges. Abbreviations: MF, medial frontal; FP, fronto-parietal; DMN, default mode; Mot, motor/sensory; VI, visual a; VII, visual b; Vas, visual assoc; SAL, salience; SC, subcortical; CBL, cerebellum/ brainstem.

To characterize the networks of loss processing, we summarized the patterns of connectivity based on the number of connections within and between canonical neural networks (Shen et al., 2013) (**Figure 2B**). By definition, positive and negative networks do not contain overlapping connections; a single edge cannot be part of both a positive and a negative network. However, positive and negative networks included connections within and between similar large-scale canonical neural networks. The positive networks included relatively more connections, involving the medial frontal, frontoparietal, and motor/sensory networks. The negative networks included relatively more connections between motor/sensory and medial frontal; between visual association and fronto-parietal; and between fronto-parietal and cerebellar networks. The positive network was further characterized by more within-network connections across medial frontal and fronto-parietal and motor/sensory and cerebellar networks, whereas the negative network included more within-network connections for motor/sensory and medial frontal networks.

#### 3.1.3 Neurotransmitters associated with network predictors of alcohol use severity: loss processing

Cross-region spatial correlation analyses revealed a significant link between the network correlates of alcohol severity PC1 and serotonergic (5-HT) and GABAergic densities (**Figure 3A**). The 5-HT system involved specifically the 5-HT1a receptors and the GABAergic system involved the GABAa2 receptors (**Figure 3B**).

**Figure 3.**
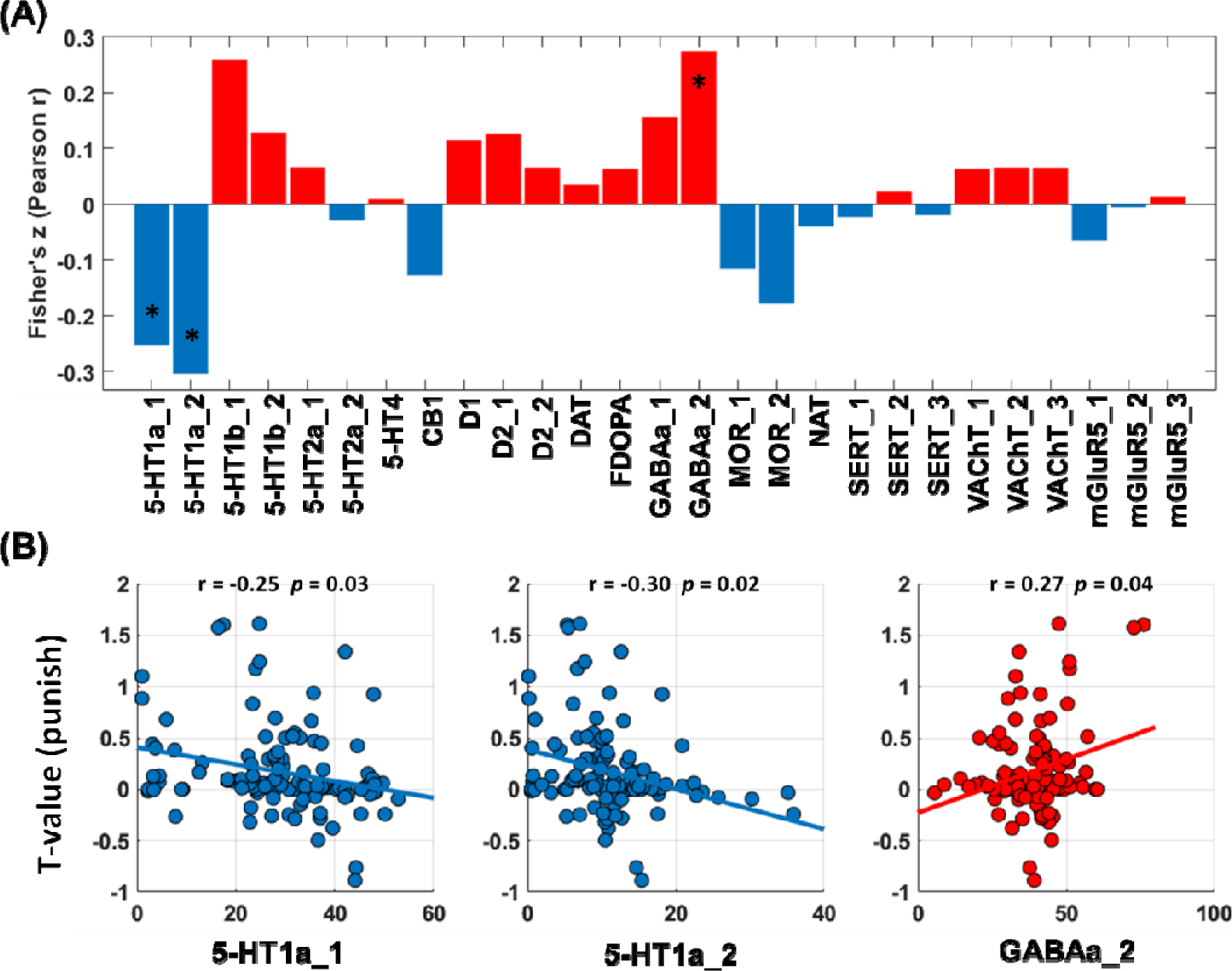
Correlations between T values of whole-brain regression of the contrast “loss-baseline” against PC1 and neurotransmitter distribution maps. **(A)** T maps and neurotransmitter correlation analysis results (red/blue each represents positive/negative Pearson’s r). **(B)** Transporter or receptor systems that are significantly associated with T maps. Abbreviations: 5-HT,5-hydroxytryptamine (serotonin); CB1, cannabinoid type 1; D, dopamine receptor; DAT, dopamine transporter; FDOPA, fluorodopa, an analog of L-DOPA to assess the nigrostriatal dopamine system; GABAa, gamma-aminobutyric acid a; MOR, mu opioid receptor; NAT, noradrenaline transporter; SERT, serotonin transporter; VAChT, vesicular acetylcholine transporter; mGluR5, metabotropic glutamate type 5.

### 3.2 Predicting drinking severity: win processing

#### 3.2.1 CPM of win processing

In CPM of win processing in the gambling task (**Figure 4A**), the overall model (positive and negative networks combined) successfully predicted drinking severity PC1 (r = 0.25, *p* = 0.001), as did connectivity within the positive (r = 0.25, *p* = 0.001) and negative (r = 0.25, *p* = 0.001) networks separately (**Figure 4B**).

**Figure 4.**
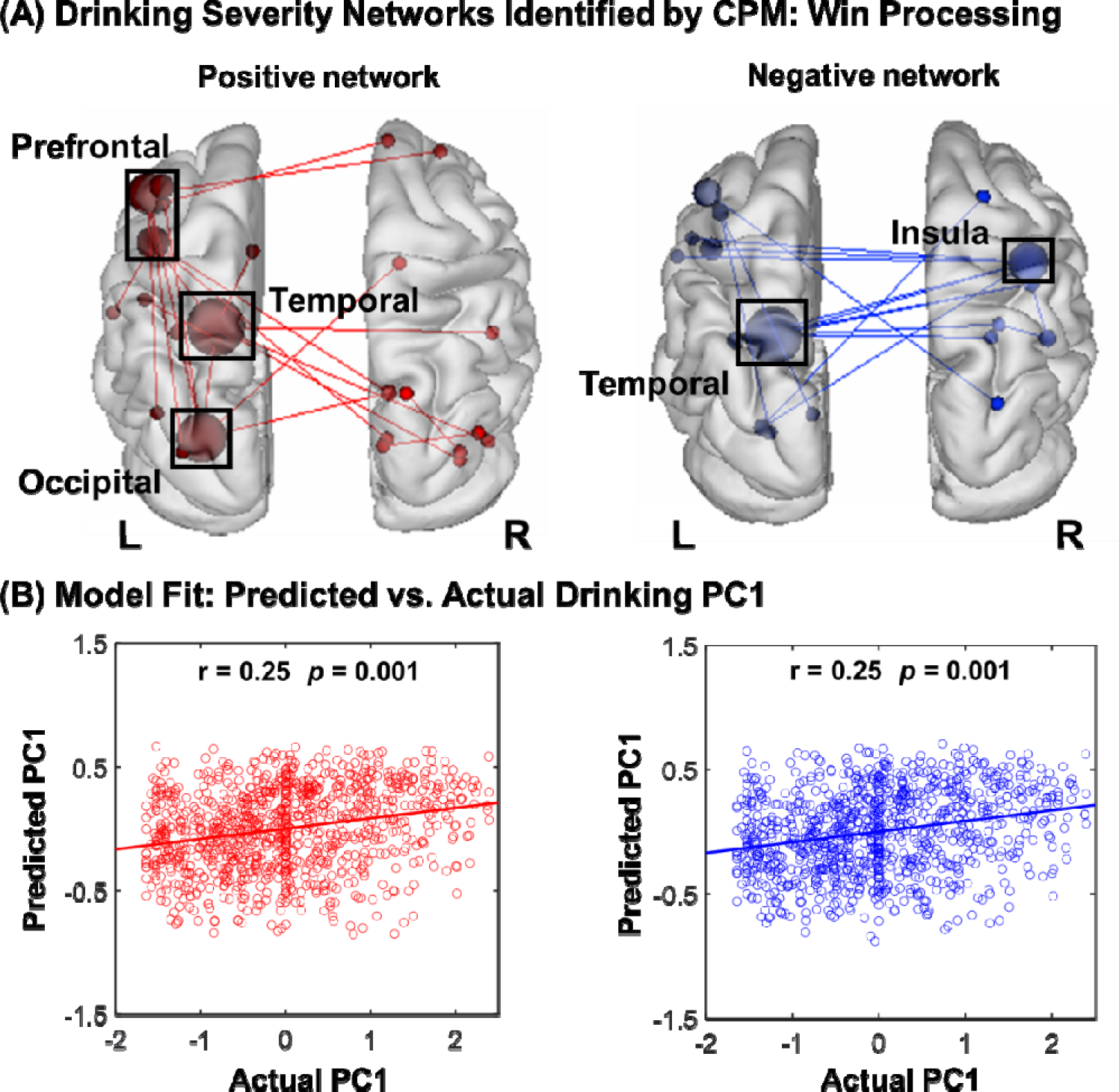
Macroscale network connectivities of win processing in predicting drinking PC1. **(A)** shows positive (red) and negative (blue) networks. For the positive network, higher edge weights (i.e., connectivity) predict more severe drinking. For the negative network, lower edge weights predict more severe drinking. Larger spheres indicate nodes with more edges, and smaller spheres indicate fewer edges. **(B)** illustrates the correlation between actual (x-axis) and predicted (y-axis) drinking severity values generated using CPM.

#### 3.2.2 Network anatomy of win processing

Network anatomies for both networks were complex and included connections between frontal, temporal, parietal, cerebellum and brainstem lobes (**Figure 5A**). However, the spatial extent of both positive and negative networks together included only 44 edges (24 positive, 20 negative), or less than 0.12% of all possible connections correlated with drinking severity PC1. Highest-degree nodes (i.e., nodes with the most connections) for the positive network included a prefrontal node with connections to temporal, occipital and limbic nodes, temporal node with connections to cerebellar, subcortical and other temporal nodes. Highest-degree nodes for the negative network also included a temporal node with connections to insula, cerebellar, brainstem nodes, temporal node with connections to insula, limbic and other temporal nodes. Both networks included short- and long-range connections. Compared to loss processing, win processing involved more functional connections of prefrontal, subcortical, and brainstem networks. All edges showing significant correlation with PC1 are shown in **Supplementary Table S2**.

**Figure 5.**
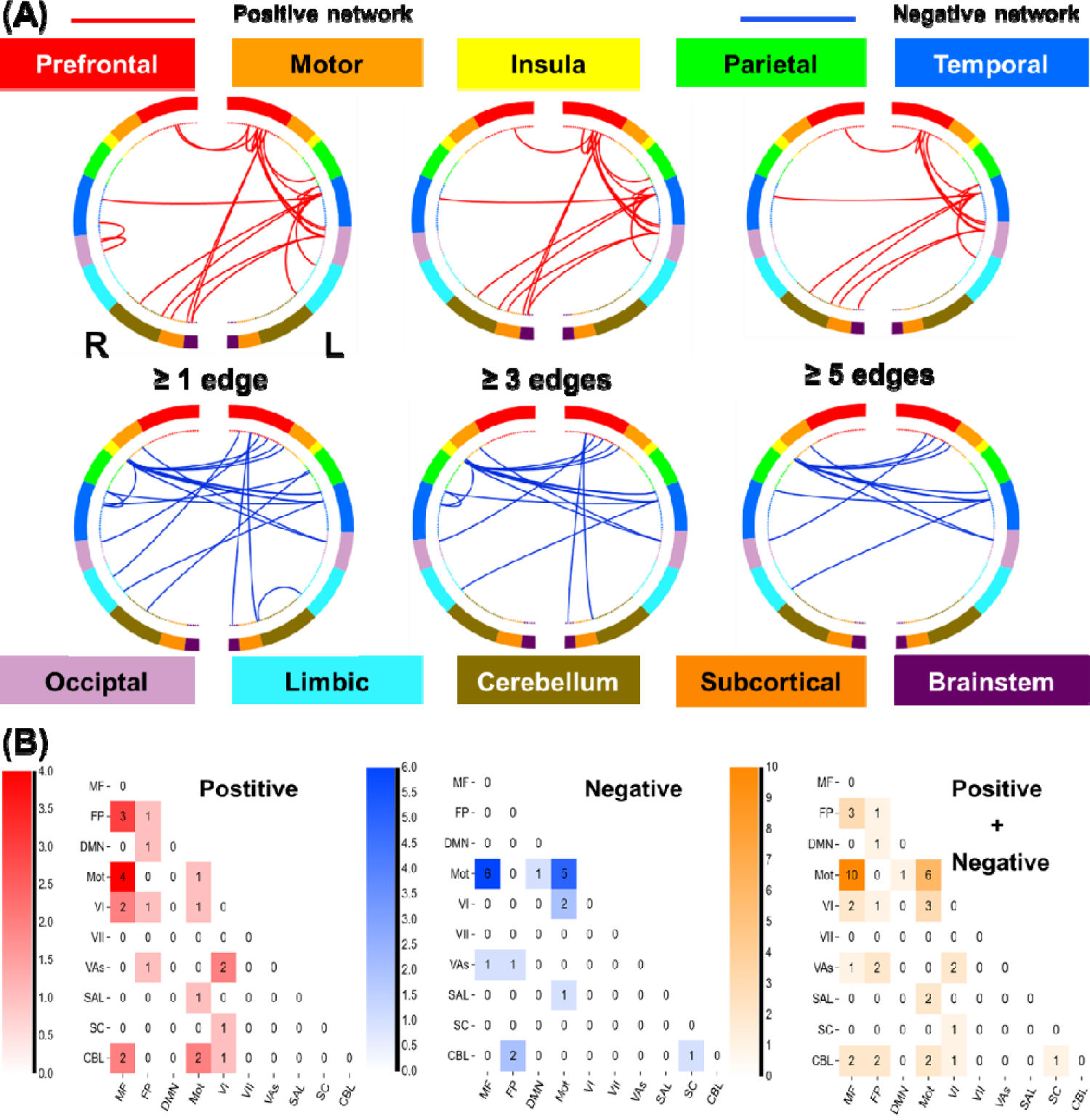
Macroscale network connectivities of win processing in predicting drinking PC1. **(A)** Positive and negative networks summarized by connectivity between macroscale brain regions during punishment blocks. From the top, brain regions are presented in approximate anatomical order, such that longer-range connections are represented by longer lines. **(B)** Positive and negative and the sum of positive and negative networks. Cells represent the total number of edges connecting nodes within and between each network, with a higher number indicating a greater number of edges. Abbreviations: MF, medial frontal; FP, frontal parietal; DMN, default mode; Mot, motor/sensory; VI, visual a; VII, visual b; Vas, visual assoc; SAL, salience; SC, subcortical; CBL, cerebellum/ brainstem.

Likewise, we summarized the connectivities based on the number of connections within and between canonical neural networks (e.g., frontoparietal, motor/sensory) for the positive and negative networks (**Figure 5B**). The positive networks included relatively more connections of medial frontal, fronto-parietal, and motor/sensory networks. The negative network included relatively more connections between motor/sensory and medial frontal. The positive network was further characterized by more within-network connections across medial frontal and fronto-parietal and motor/sensory and cerebellar networks, whereas the negative network included more within-network connections for motor/sensory and medial frontal networks.

#### 3.2.3 Neurotransmitters associated with network predictors of alcohol use severity: win processing

As with loss processing, cross-region spatial correlation analyses revealed a significant link between the network correlates of alcohol severity PC1 and serotonergic (5-HT) and GABAergic densities during win processing (**Figure 6A**). The 5-HT system involved specifically the 5-HT1a receptors and the GABAergic system involved the GABAa2 receptors (**Figure 6B**).

**Figure 6.**
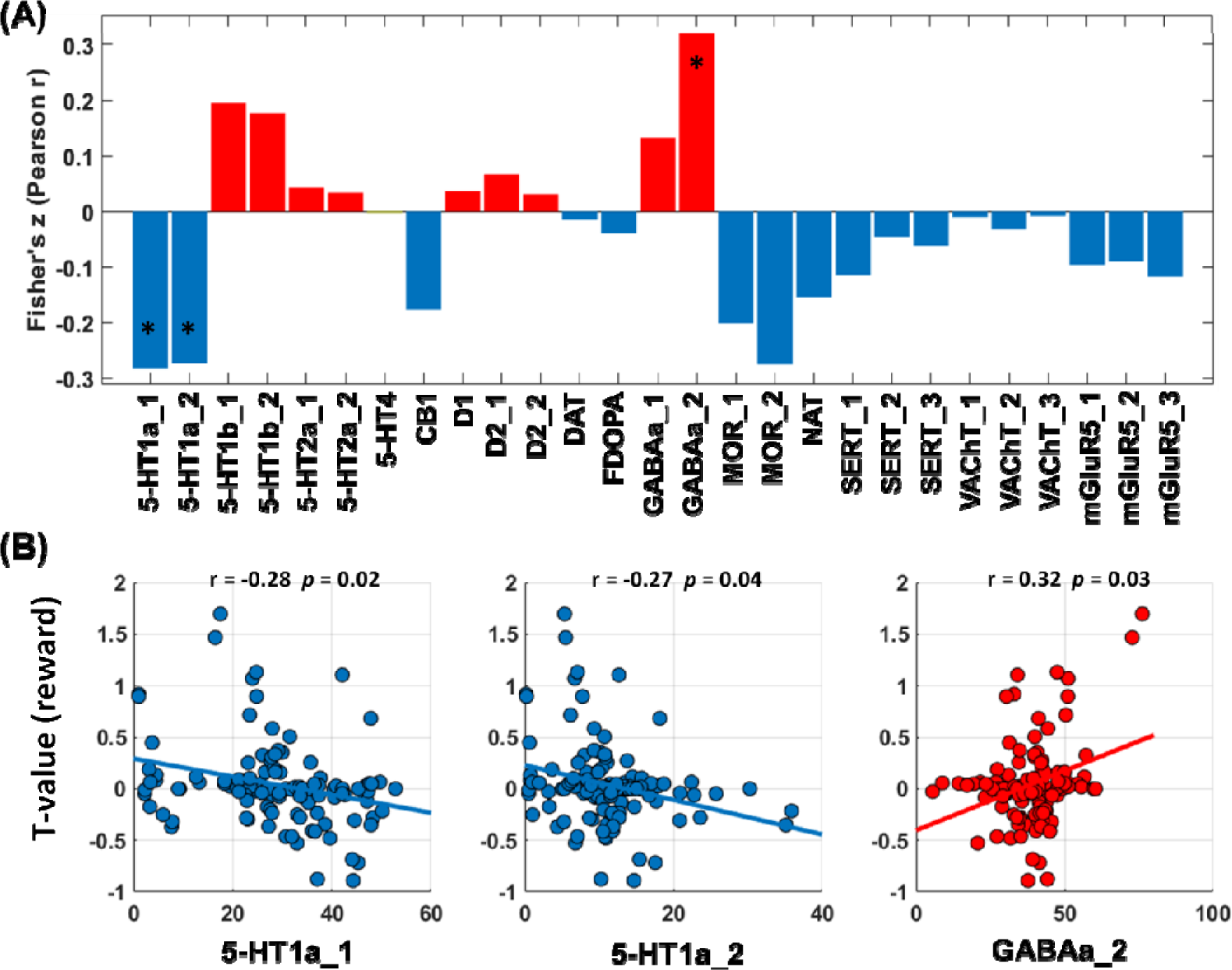
Correlations between T values of whole-brain regression of the contrast “win-baseline” against PC1 and neurotransmitter distribution maps. **(A)** T maps and neurotransmitter correlation analysis results (red/blue each represents positive/negative Pearson’s r). **(B)** Transporter or receptor systems that are significantly associated with T maps. Abbreviations: 5-HT,5-hydroxytryptamine (serotonin); CB1, cannabinoid type 1; D, dopamine receptor; DAT, dopamine transporter; FDOPA, fluorodopa, an analog of L-DOPA to assess the nigrostriatal dopamine system; GABAa, gamma-aminobutyric acid a; MOR, mu opioid receptor; NAT, noradrenaline transporter; SERT, serotonin transporter; VAChT, vesicular acetylcholine transporter; mGluR5, metabotropic glutamate type 5.

## 4. Discussion

In this study, we demonstrated the utility of a connectome-based machine learning approach in predicting alcohol use severity using whole-brain functional connectivities in a gambling task. The connectomics predominantly involving medial frontal, motor/sensory, and cerebellum/brainstem networks during punishment processing and those involving medial frontal and fronto-parietal, motor/sensory networks during reward processing predicted drinking severity. Further, with JuSpace, we demonstrated in spatial correlation analyses that these networks were associated specifically with 5-HT1a and GABAa signaling. Together, these findings highlight connectivity markers of alcohol use severity and the molecular profiles of these markers in young adults.

### 4.1 Connectivity features that predict drinking severity

With connectivity features of both loss and win processing during gambling, CPM successfully predicted drinking severity. Although the specific features varied, the positive networks included connections amongst the medial frontal, motor/sensory, and fronto-parietal networks, and the negative networks included connections between motor/sensory and medial frontal; between visual association and fronto-parietal; and between fronto-parietal and cerebellar networks for both loss and win processing. In contrast, whereas loss processing involved positive temporal, prefrontal, subcortical network connections and negative temporal and limbic, including insula, connections, win processing involved positive network connections between temporal and limbic, between prefrontal and occipital and negative network connections amongst insula, prefrontal, and temporal cortices.

These findings are broadly consistent with reports of dysfunctional activation of the network nodes during psychological processes of importance to habitual drinking. For instance, an earlier work associated duration of alcohol use with lower activation in the right inferior frontal gyrus extending to superior temporal gyrus during inhibitory control, which was not observed for age-related changes in nondrinkers (Hu et al., 2016). Another study demonstrated cue-craving circuits that involved connectivities with the frontal, parietal, and temporal brain regions in AUD participants vs. controls (Hornoiu et al., 2023). Whereas it is challenging to relate these activity or connectivity findings directly to the connectomics features identified in the current study, it appears that the same brain regions may participate in various psychological processes that conduce to alcohol consumption via their wide network connectivities.

On the other hand, these connectivity results contrasted with our earlier findings that loss but not win reactivities distinguished individual variation in drinking (Li et al., 2022) and suggested connectivity features as additional neural markers of alcohol use severity. Secondly, shared connectivity features of loss and win processing suggest the potential roles of saliency circuit dysfunction in alcohol misuse. The current findings can also be discussed with earlier CPM studies of drinking or other substance use. For instance, Rapuano and colleagues demonstrated that both resting and reward-related connectomics in developing brains may predict risk for substance use, as reflected in e.g., substance-related behavioral measures and family history of drug use (Rapuano et al., 2021). Another study applied CPM to the International Neuroimaging Data-sharing Initiative database and identified functional connectivity from the DMN to the sensorimotor, opercular, and occipital network in predicting empathy in healthy subjects but not in abstinent drinkers, suggesting individual variation in the degree and pattern of network disruption as a result of chronic alcohol exposure (Yao et al., 2022). Other studies of CPM reported connectivities of the motor/sensory, salience, and executive control networks during cue exposure in prediction of dependence severity in male smokers (Lin et al., 2022) and of stronger motor/sensory within-network connectivity, and reduced connectivity between the motor/sensory and medial frontal, default mode, and frontoparietal networks in distinguishing opioid from other substance use disorders (Lichenstein et al., 2021).

An important question concerns whether the connectomics features would predict future drinking, as was successfully demonstrated for reward-related fMRI data in a treatment study of cocaine use disorders (Yip et al., 2019). In a magnetoencephalography (MEG) study of adolescents described the relationship between functional Connectivity (FC) during an inhibitory control (IC) task and development of heavy drinking over a two-year period. The results showed that higher beta-band FC in the prefrontal and temporal regions at baseline predicted higher levels of future alcohol consumption. Further, greater future alcohol consumption was associated with more severe reduction in the same FC’s (Anton-Toro et al., 2023). Another study showed that greater severity of binge drinking in college students was negatively associated with connectivity between the DMN and ventral attention network, although CPM failed to identify a generalizable predictive model of longitudinal changes in connectivity edges over follow-up of two years (Tong et al., 2021). It remains to be seen whether the current network markers predict future alcohol use and identify individuals at risk for AUD.

### 4.2 Molecular profiles of the connectivity networks

We examined the molecular profiles of the network markers, and the results highlighted the role of the GABAergic and serotonergic signaling in alcohol use severity. This finding is consistent with a large body of clinical and preclinical research implicating the GABAergic and serotonergic systems in alcohol misuse (Davies, 2003; Maccioni and Colombo, 2019; Andersen et al., 2021; Logge et al., 2022).

Alcohol mimics the activity of GABA by binding to GABA receptors and inhibits neuronal activities, leading to widespread suppression of brain function. Other studies implicated the GABAergic systems in dependent drinking. For instance, injection of muscimol, a GABAa receptor agonist, in the ventral tegmental area elevated voluntary drinking in alcohol-preferring AA rats (Dudek and Hyytia, 2016). In a study quantifying the levels of transcript expression of GABAa receptor mRNA in postmortem brain tissues, those with a diagnosis of AUD who died of cirrhotic liver disease (suggesting higher level of alcohol use) as compared to controls has significantly higher expression of the transcripts in dorsolateral prefrontal and primary motor cortices (Ashton et al., 2022).

Preclinical work demonstrated the roles of 5-HT1A-dependent regulation of binge drinking from short-to longer-term alcohol exposure that involves the dentate gyrus of the hippocampus (Belmer et al., 2022). In a drinking-in-the-dark (DID) paradigm to model chronic binge-like voluntary alcohol consumption in mice, selective partial activation of 5-HT1A receptors by tandospirone (5-HT1A partial agonist) prevented alcohol withdrawal-induced anxiety-related behavior and binge-like ethanol intake. Further, DID-elicited deficits in neurogenesis in the dorsal hippocampus were reversed by chronic treatment with tandospirone (Belmer et al., 2018). An earlier study characterized the opposing effects of stimulation of somatodendritic 5-HT1A receptors at lower doses and postsynaptic 5-HT1A receptors at higher doses on alcohol intake in Long-Evans rats (McKenzie-Quirk and Miczek, 2003). In PET imaging of 5-HT1A receptor binding in alcohol-naïve rhesus, the binding potential increased in the raphe nuclei (vs. the cerebellum as a reference region) after chronic ethanol self-administration. Further, baseline 5-HT1A binding in the raphe nuclei showed a positive correlation with average daily ethanol self-administration (Hillmer et al., 2014). In a large sample of adolescents, 5-HTTLPR low-activity allele carriers exposed to higher levels of family conflict were more likely to engage in alcohol misuse than non-carriers (Kim et al., 2020).

Together, the current findings are consistent with this previous body of work implicating GABAergic and serotonergic signaling in alcohol misuse. However, many other studies implicated the glutamatergic (Morris et al., 1986; Bliss and Collingridge, 1993; White et al., 2000), dopaminergic (Travis E. Baker et al., 2019), cannabinoid (Ridge et al., 2009), and GABAb (Marron Fernandez de Velasco et al., 2023) signaling in alcohol misuse. Some of these systems which have yet to be covered by the JuSpace toolbox need to be further investigated along with the network markers of alcohol use severity.

### 4.3 Limitations and conclusions

A few limitations need to be considered for this study. First, the networks identified from CPM included nodes from Shen’s atlas of 268 ROIs, whereas the neurotransmitter maps were of 119 brain regions. This discrepancy may have accounted for the missing links with other neurotransmitter systems. Second, the HCP data represent a non-clinical sample; it thus remains to be seen whether the current findings can be generalized to alcohol use disorders. Third, alcohol misuse involves many other comorbidities, including smoking and internet gaming disorder (Zhou et al., 2022) as well as depression and anxiety (Amanda L. Baker et al., 2012; Debell et al., 2014) that may implicate changes in cerebral connectomics. Although a recent study of a large cohort of adolescents highlighted resting-state connectivity features across the majority of networks more strongly predictive of drinking than smoking (Gazula et al., 2023), it remains to be seen whether the connectivity features identified here are specific to alcohol use severity. Finally, many large-scale studies have characterized the gray matter volumetric and thickness and white matter integrity markers of alcohol misuse (Galinowski et al., 2020; Harper et al., 2021; Logtenberg et al., 2022; Rane et al., 2022). Future studies can examine how functional and structural markers compare in the effect size of prediction and whether the combination of multimodal imaging makers improves the performance of CPM.

In conclusion, this study demonstrates that patterns of whole-brain connectivity during loss and win processing can predict drinking severity and these connectivity markers are associated with the serotonergic and GABAa signaling. These findings demonstrate that individual differences in connectivity within large-scale neural networks as implicated in punishment and reward responses contribute to the severity of alcohol misuse outcomes. As such, these “neural fingerprints” may represent appropriate targets for future intervention efforts.

## Authors Contributions

Author CS L and YL (Youjun Liu) designed the study and wrote the protocol. Author GL, Y Y-L and YL (Yashuang Li) performed the literature search and data analyses. Authors BL, YC, DH and LY assisted in statistical analysis, and author GL and YL (Yashuang Li) wrote the first draft of the manuscript. All authors contributed to revisions and have approved the final version of the manuscript. All authors agree to be accountable for all aspects of the work in ensuring that questions related to the accuracy or integrity of any part of the work are appropriately investigated and resolved

## Funding Support and Acknowledgement

The current study is supported by National Key Research and Development Program of China (2021YFA1000200, 2021YFA1000202), Beijing Nova Program (20230484469), National Natural Science Foundation of China (U20A20388), China Postdoctoral Science Foundation (2022M720326) and NIH grant (DA051922, C-SRL). Data were provided by the Human Connectome Project, WU-Minn Consortium (Principal Investigators: David Van Essen and Kamil Ugurbil; 1U54MH091657) funded by the 16 NIH Institutes and Centers that support the NIH Blueprint for Neuroscience Research; and by the McDonnell Center for Systems Neuroscience at Washington University.

## Data Availability

All data needed to evaluate the conclusions in the paper are shown in the paper and the supplement. All raw data supporting the analyses and findings of this study are available from HCP.

## Supporting information

Supplemental Results

## Data Availability

https://db.humanconnectome.org/app/template/Login.vm;jsessionid=E37EF29E03161C15E526A149C8554B60

## References

Andersen KAA, Carhart-Harris R, Nutt DJ, Erritzoe D (2021) Therapeutic effects of classic serotonergic psychedelics: A systematic review of modern-era clinical studies. Acta psychiatrica Scandinavica 143:101–118.

Anton-Toro LF, Shpakivska-Bilan D, Del Cerro-Leon A, Bruna R, Uceta M, Garcia-Moreno LM, Maestu F (2023) Longitudinal change of inhibitory control functional connectivity associated with the development of heavy alcohol drinking. FRONT PSYCHOL 14:1069990–1069990.

Antons S, Yip SW, Lacadie CM, Dadashkarimi J, Scheinost D, Brand M, Potenza MN (2023) Connectome-based prediction of craving in gambling disorder and cocaine use disorder. DIALOGUES CLIN NEURO 25:33–42.

Ashton MK, Rueda AVL, Ho AM, Noor Aizin N, Sharma H, Dodd PR, Stadlin A, Camarini R (2022) Sex differences in GABA(A) receptor subunit transcript expression are mediated by genotype in subjects with alcohol-related cirrhosis of the liver. Genes Brain Behav 21:e12785.

Baker AL, Thornton LK, Hiles S, Hides L, Lubman DI (2012) Psychological interventions for alcohol misuse among people with co-occurring depression or anxiety disorders: A systematic review. J AFFECT DISORDERS 139:217–229.

Baker TE et al. (2019) Modulation of orbitofrontal-striatal reward activity by dopaminergic functional polymorphisms contributes to a predisposition to alcohol misuse in early adolescence. PSYCHOL MED 49:801–810.

Barch DM et al. (2013) Function in the human connectome: task-fMRI and individual differences in behavior. Neuroimage 80:169–189.

Beaty RE, Kenett YN, Christensen AP, Rosenberg MD, Benedek M, Chen Q, Fink A, Qiu J, Kwapil TR, Kane MJ, Silvia PJ (2018) Robust prediction of individual creative ability from brain functional connectivity. Proceedings of the National Academy of Sciences 115:1087–1092.

Belmer A, Patkar OL, Lanoue V, Bartlett SE (2018) 5-HT1A receptor-dependent modulation of emotional and neurogenic deficits elicited by prolonged consumption of alcohol. SCI REP-UK 8:2099–2012.

Belmer A, Depoortere R, Beecher K, Newman-Tancredi A, Bartlett SE (2022) Neural serotonergic circuits for controlling long-term voluntary alcohol consumption in mice. MOL PSYCHIATR 27:4599–4610.

Bliss TV, Collingridge GL (1993) A synaptic model of memory: long-term potentiation in the hippocampus. Nature 361:31–39.

Chen J, Wei Y, Xue K, Han S, Wang C, Wen B, Cheng J (2023) The interaction between first-episode drug-naïve schizophrenia and age based on gray matter volume and its molecular analysis: a multimodal magnetic resonance imaging study. Psychopharmacology 240:813–826.

Chen Y, Li C-SR (2023) Appetitive and aversive cue reactivities differentiate neural subtypes of alcohol drinkers. Addiction Neuroscience 7:100089.

Cheng W, Rolls ET, Robbins TW, Gong W, Liu Z, Lv W, Du J, Wen H, Ma L, Quinlan EB, Garavan H, Artiges E, Papadopoulos Orfanos D, Smolka MN, Schumann G, Kendrick K, Feng J (2019) Decreased brain connectivity in smoking contrasts with increased connectivity in drinking. eLife 8:e40765.

Cui S, Jiang P, Cheng Y, Cai H, Zhu J, Yu Y (2023) Molecular mechanisms underlying resting-state brain functional correlates of behavioral inhibition. Neuroimage 283:120415.

Davies M (2003) The role of GABAA receptors in mediating the effects of alcohol in the central nervous system. Journal of psychiatry & neuroscience : JPN 28:263–274.

Debell F, Fear NT, Head M, Batt-Rawden S, Greenberg N, Wessely S, Goodwin L (2014) A systematic review of the comorbidity between PTSD and alcohol misuse. Soc Psychiatry Psychiatr Epidemiol 49:1401–1425.

Dudek M, Hyytia P (2016) Alcohol preference and consumption are controlled by the caudal linear nucleus in alcohol-preferring rats. Eur J Neurosci 43:1440–1448.

Dugre JR, Potvin S (2022) The origins of evil: From lesions to the functional architecture of the antisocial brain. FRONT PSYCHIATRY 13:969206–969206.

Dukart J, Holiga S, Rullmann M, Lanzenberger R, Hawkins PCT, Mehta MA, Hesse S, Barthel H, Sabri O, Jech R, Eickhoff SB (2021) JuSpace: A tool for spatial correlation analyses of magnetic resonance imaging data with nuclear imaging derived neurotransmitter maps. Hum Brain Mapp 42:555–566.

Feng C, Yuan J, Geng H, Gu R, Zhou H, Wu X, Luo Y (2018) Individualized prediction of trait narcissism from whole-brain resting-state functional connectivity. Hum Brain Mapp 39:3701–3712.

Finn ES, Shen X, Scheinost D, Rosenberg MD, Huang J, Chun MM, Papademetris X, Constable RT (2015) Functional connectome fingerprinting: identifying individuals using patterns of brain connectivity. Nat Neurosci 18:1664–1671.

Fiore A, Preziosa P, Tedone N, Margoni M, Vizzino C, Mistri D, Gueye M, Rocca MA, Filippi M (2023) Correspondence among gray matter atrophy and atlas-based neurotransmitter maps is clinically relevant in multiple sclerosis. MOL PSYCHIATR 28:1770–1782.

Galinowski A et al. (2020) Heavy drinking in adolescents is associated with change in brainstem microstructure and reward sensitivity. ADDICT BIOL 25:e12781-n/a.

Gazula H et al. (2023) Federated Analysis in COINSTAC Reveals Functional Network Connectivity and Spectral Links to Smoking and Alcohol Consumption in Nearly 2,000 Adolescent Brains. Neuroinform 21:287–301.

Harper J, Malone SM, Wilson S, Hunt RH, Thomas KM, Iacono WG (2021) The Effects of Alcohol and Cannabis Use on the Cortical Thickness of Cognitive Control and Salience Brain Networks in Emerging Adulthood: A Co-twin Control Study. BIOL PSYCHIAT 89:1012–1022.

Hillmer AT, Wooten DW, Tudorascu DL, Barnhart TE, Ahlers EO, Resch LM, Larson JA, Converse AK, Moore CF, Schneider ML, Christian BT (2014) The effects of chronic alcohol self-administration on serotonin-1A receptor binding in nonhuman primates. DRUG ALCOHOL DEPEN 144:119–126.

Hirjak D, Schmitgen MM, Werler F, Wittemann M, Kubera KM, Wolf ND, Sambataro F, Calhoun VD, Reith W, Wolf RC (2022) Multimodal MRI data fusion reveals distinct structural, functional and neurochemical correlates of heavy cannabis use. ADDICT BIOL 27:e13113-n/a.

Hornoiu IL, Lee AM, Tan H, Nakovics H, Bach P, Mann K, Kiefer F, Sommer WH, Vollstädt-Klein S (2023) The Role of Unawareness, Volition, and Neural Hyperconnectivity in Alcohol Use Disorder: A Functional Magnetic Resonance Imaging Study. BIOL PSYCHIAT-COGN N 8:660–671.

Hu S, Zhang S, Chao HH, Krystal JH, Li C-SR (2016) Association of Drinking Problems and Duration of Alcohol Use to Inhibitory Control in Nondependent Young Adult Social Drinkers. ALCOHOL CLIN EXP RES 40:319–328.

Ide JS, Li HT, Chen Y, Le TM, Li CSP, Zhornitsky S, Li CR (2020) Gray matter volumetric correlates of behavioral activation and inhibition system traits in children: An exploratory voxel-based morphometry study of the ABCD project data. Neuroimage 220:117085.

Kahn RE, Chiu PH, Deater-Deckard K, Hochgraf AK, King-Casas B, Kim-Spoon J (2018) The Interaction Between Punishment Sensitivity and Effortful Control for Emerging Adults’ Substance Use Behaviors. Subst Use Misuse 53:1299–1310.

Kim J, Zaso MJ, Desalu JM, Park A (2020) Interaction between the 5-hydroxytryptamine transporter-linked polymorphic region (5-HTTLPR) and negative life events in adolescent heavy drinking. J STUD ALCOHOL DRUGS 81:566–574.

Koob GF, Volkow ND (2016) Neurobiology of addiction: a neurocircuitry analysis. The lancet Psychiatry 3:760–773.

Le TM, Zhornitsky S, Wang W, Ide J, Zhang S, Li CR (2019) Posterior Cingulate Cortical Response to Active Avoidance Mediates the Relationship between Punishment Sensitivity and Problem Drinking. J Neurosci 39:6354–6364.

Li G, Chen Y, Chaudhary S, Tang X, Li C-SR (2022) Loss and frontal striatal reactivities characterize alcohol use severity and rule-breaking behavior in young adult drinkers. Biological Psychiatry: Cognitive Neuroscience and Neuroimaging.

Li G, Chen Y, Wang W, Dhingra I, Zhornitsky S, Tang X, Li C-SR (2020) Sex Differences in Neural Responses to the Perception of Social Interactions. Front Hum Neurosci 14:565132–565132.

Li G, Chen Y, Le TM, Zhornitsky S, Wang W, Dhingra I, Zhang S, Tang X, Li C-SR (2021a) Perceived friendship and binge drinking in young adults: A study of the Human Connectome Project data. Drug and Alcohol Dependence 224:108731.

Li G, Le TM, Wang W, Zhornitsky S, Chen Y, Chaudhary S, Zhu T, Zhang S, Bi J, Tang X, Li C-SR (2021b) Perceived stress, self-efficacy, and the cerebral morphometric markers in binge-drinking young adults. NeuroImage: Clinical 32:102866.

Lichenstein SD, Scheinost D, Potenza MN, Carroll KM, Yip SW (2021) Dissociable neural substrates of opioid and cocaine use identified via connectome-based modelling. MOL PSYCHIATR 26:4383–4393.

Lin X, Zhu X, Zhou W, Zhang Z, Li P, Dong G, Meng S, Deng J, Lu L (2022) Connectome-based predictive modelling of smoking severity in smokers. Addict Biol 27:e13242.

Logge WB, Morley KC, Haber PS (2022) GABA(B) Receptors and Alcohol Use Disorders: Clinical Studies. Current topics in behavioral neurosciences 52:195–212.

Logtenberg E, Overbeek MF, Pasman JA, Abdellaoui A, Luijten M, van Holst RJ, Vink JM, Denys D, Medland SE, Verweij KJH, Treur JL (2022) Investigating the causal nature of the relationship of subcortical brain volume with smoking and alcohol use. BRIT J PSYCHIAT 221:377–385.

Maccioni P, Colombo G (2019) Potential of GABA(B) Receptor Positive Allosteric Modulators in the Treatment of Alcohol Use Disorder. CNS drugs 33:107–123.

Marron Fernandez de Velasco E, Tipps ME, Haider B, Souders A, Aguado C, Rose TR, Vo BN, DeBaker MC, Luján R, Wickman K (2023) Ethanol-Induced Suppression of G Protein– Gated Inwardly Rectifying K+–Dependent Signaling in the Basal Amygdala. Biol Psychiatry 94:863–874.

McKenzie-Quirk SD, Miczek KA (2003) 5-HT1A agonists: Alcohol drinking in rats and squirrel monkeys. PSYCHOPHARMACOLOGY 167:145–152.

Moreno Padilla M, O’Halloran L, Bennett M, Cao Z, Whelan R (2017) Impulsivity and Reward Processing Endophenotypes in Youth Alcohol Misuse. Current Addiction Reports 4:350–363.

Morris RG, Anderson E, Lynch GS, Baudry M (1986) Selective impairment of learning and blockade of long-term potentiation by an N-methyl-D-aspartate receptor antagonist, AP5. Nature 319:774–776.

Mummaneni A, Kardan O, Stier AJ, Chamberlain TA, Chao AF, Berman MG, Rosenberg MD (2023) Functional brain connectivity predicts sleep duration in youth and adults. Human brain mapping.

Peng Z, Dai C, Cai X, Zeng L, Li J, Xie S, Wang H, Yang T, Shao Y, Wang Y (2020) Total Sleep Deprivation Impairs Lateralization of Spatial Working Memory in Young Men. Frontiers in neuroscience 14:562035.

Pengo M et al. (2023) Early neurotransmitters changes in prodromal frontotemporal dementia: A GENFI study. NEUROBIOL DIS 179:106068–106068.

Premi E, Dukart J, Mattioli I, Libri I, Pengo M, Gadola Y, Cotelli M, Manenti R, Binetti G, Gazzina S, Alberici A, Magoni M, Koch G, Gasparotti R, Padovani A, Borroni B (2023) Unravelling neurotransmitters impairment in primary progressive aphasias. HUM BRAIN MAPP 44:2245–2253.

Rane RP et al. (2022) Structural differences in adolescent brainscan predict alcohol misuse. ELIFE 11.

Rapuano KM, Rosenberg MD, Maza MT, Dennis NJ, Dorji M, Greene AS, Horien C, Scheinost D, Todd Constable R, Casey BJ (2021) Corrigendum to “Behavioral and brain signatures of substance use vulnerability in childhood” [Developmental Cognitive Neuroscience 46 (December) (2020) 100878]. Dev Cogn Neurosci 47:100891.

Ren J, Yan L, Zhou H, Pan C, Xue C, Wu J, Liu W (2023) Unraveling neurotransmitter changes in de novo GBA-related and idiopathic Parkinson’s disease. NEUROBIOL DIS 185:106254–106254.

Ridge JP, Ho AMC, Dodd PR (2009) Sex Differences in NMDA Receptor Expression in Human Alcoholics. Alcalc 44:594–601.

Rosenberg MD, Finn ES, Scheinost D, Papademetris X, Shen X, Constable RT, Chun MM (2016) A neuromarker of sustained attention from whole-brain functional connectivity. Nat Neurosci 19:165–171.

Shen X, Tokoglu F, Papademetris X, Constable RT (2013) Groupwise whole-brain parcellation from resting-state fMRI data for network node identification. NEUROIMAGE 82:403–415.

Shen X, Finn ES, Scheinost D, Rosenberg MD, Chun MM, Papademetris X, Constable RT (2017) Using connectome-based predictive modeling to predict individual behavior from brain connectivity. Nature protocols 12:506–518.

Tang C, Ren P, Ma K, Li S, Wang X, Guan Y, Zhou J, Li T, Liang X, Luan G (2022) The correspondence between morphometric MRI and metabolic profile in Rasmussen ’ s encephalitis. NEUROIMAGE-CLIN 33:102918–102918.

Tong TT, Vaidya JG, Kramer JR, Kuperman S, Langbehn DR, O’Leary DS (2021) Impact of binge drinking during college on resting state functional connectivity. DRUG ALCOHOL DEPEN 227:108935–108935.

Van Essen DC et al. (2012) The Human Connectome Project: A data acquisition perspective. NeuroImage 62:2222–2231.

White AM, Matthews DB, Best PJ (2000) Ethanol, memory, and hippocampal function: a review of recent findings. Hippocampus 10:88–93.

Wu H, Zhou C, Guan X, Bai X, Guo T, Wu J, Chen J, Wen J, Wu C, Cao Z, Liu X, Gao T, Gu L, Huang P, Xu X, Zhang B, Zhang M (2023) Functional connectomes of akinetic-rigid and tremor within drug-naïve Parkinson’s disease. CNS NEUROSCI THER 29:3507–3517.

Yao G, Wei L, Jiang T, Dong H, Baeken C, Wu GR (2022) Neural mechanisms underlying empathy during alcohol abstinence: evidence from connectome-based predictive modeling. Brain Imaging Behav 16:2477–2486.

Yip SW, Scheinost D, Potenza MN, Carroll KM (2019) Connectome-Based Prediction of Cocaine Abstinence. Am J Psychiatry 176:156–164.

Zhou WR, Wang YM, Wang M, Wang ZL, Zheng H, Wang MJ, Potenza MN, Dong GH (2022) Connectome-based prediction of craving for gaming in internet gaming disorder. Addict Biol 27:e13076.

