## Supplemental Results for "Functional networks of reward and punishment processing and their molecular profiles predicting the severity of young adult drinking"

**Supplementary Methods**

*Imaging protocol, behavioral tasks, and data preprocessing*

MRI was done using a customized 3 T Siemens Connectome Skyra with a standard 32-channel Siemens receiver head coil and a body transmission coil. T1-weighted high-resolution structural images were acquired using a 3D MPRAGE sequence with 0.7 mm isotropic resolution (FOV = 224 × 224 mm, matrix = 320 × 320, 256 sagittal slices, TR = 2400 ms, TE = 2.14 ms, TI = 1000 ms, FA = 8°) and used to register functional MRI data to a standard brain space. FMRI data were collected using gradient-echo echo-planar imaging (EPI) with 2.0 mm isotropic resolution (FOV = 208 × 180 mm, matrix = 104 × 90, 72 slices, TR = 720 ms, TE = 33.1 ms, FA = 52°, multi-band factor = 8).

Participants completed two runs of a gambling task each with 4 blocks (~3 m and 12 s each run) – 2 of punishment and 2 of reward – in a fixed order (run 1: punishment – reward – punishment – reward; and run 2: reward – punishment – punishment – reward) with a fixation period (15 s) between blocks. The participants guessed whether the number of a mystery card (represented by a ‘?’ and ranging from 1 to 9) was larger or smaller than 5 by pressing a corresponding button (Barch et al., 2013). The feedbacks comprised a green up-pointing arrow for correct guess and $1 win, a red down-pointing arrow for $0.5 loss; or a gray double-headed arrow for a wash (mystery card number = 5). The mystery number was controlled by the program and shown for 1.5 s, followed by the feedback for 1.0 s. There was a 1.0 s inter-trial interval with a “+” shown on the screen. Each block contained 8 trials. In reward blocks, 6 win trials were pseudo-randomly interleaved with either 1 neutral and 1 loss trial, 2 neutral trials, or 2 loss trials. In punishment blocks, 6 loss trials were interleaved with either 1 neutral and 1 win trial, 2 neutral trials, or 2 win trials. Thus, the amount of money won was the same across subjects.

BOLD data were analyzed with Statistical Parametric Mapping (SPM8, Welcome Department of Imaging Neuroscience, University College London, U.K.), following our published routines (Wang et al., 2020; Zhang et al., 2019; Zhornitsky et al., 2019). Images of each individual subject were first realigned (motion corrected). A mean functional image volume was constructed for each subject per run from the realigned image volumes. These mean images were co-registered with the high-resolution structural MPRAGE image and then segmented for normalization with affine registration followed by nonlinear transformation. The normalization parameters determined for the structural volume were then applied to the corresponding functional image volumes for each subject. The voxel is of 2x2x2 mm^3^ after spatial normalization. Finally, the images were smoothed with a Gaussian kernel of 4 mm at Full Width at Half Maximum.

*The GLM and 2^nd^-level analyses*

Briefly, a statistical analytical block design was constructed for each individual subject, using a general linear model (GLM) by convolving the canonical hemodynamic response function (HRF) with a boxcar function in SPM. Realignment parameters in all six dimensions were entered in the model as covariates. We constructed for each individual subject the statistical contrast “reward vs. baseline” and “punishment vs. baseline”, with baseline = 15-s fixation period between blocks in gambling task.

**Supplementary References**

Barch, D.M., Burgess, G.C., Harms, M.P., Petersen, S.E., Schlaggar, B.L., Corbetta, M., Glasser, M.F., Curtiss, S., Dixit, S., Feldt, C., Nolan, D., Bryant, E., Hartley, T., Footer, O., Bjork, J.M., Poldrack, R., Smith, S., Johansen-Berg, H., Snyder, A.Z., Van Essen, D.C., Consortium, W.U.-M.H., 2013. Function in the human connectome: task-fMRI and individual differences in behavior. Neuroimage 80, 169-189.

Wang, W., Zhornitsky, S., Le, T.M., Zhang, S., Li, C.-S.R., 2020. Heart Rate Variability, Cue-Evoked Ventromedial Prefrontal Cortical Response, and Problem Alcohol Use in Adult Drinkers. Biological Psychiatry: Cognitive Neuroscience and Neuroimaging 5, 619-628.

Zhang, S., Zhornitsky, S., Le, T.M., Li, C.R., 2019. Hypothalamic Responses to Cocaine and Food Cues in Individuals with Cocaine Dependence. Int J Neuropsychopharmacol 22, 754-764.

Zhornitsky, S., Zhang, S., Ide, J.S., Chao, H.H., Wang, W., Le, T.M., Leeman, R.F., Bi, J., Krystal, J.H., Li, C.R., 2019. Alcohol Expectancy and Cerebral Responses to Cue-Elicited Craving in Adult Nondependent Drinkers. Biol Psychiatry Cogn Neurosci Neuroimaging 4, 493-504.
